# Assessment of respiratory droplet transmission during the ophthalmic slit lamp exam: a particle tracking analysis

**DOI:** 10.1101/2020.06.25.20140335

**Authors:** Sahil H. Shah, Anupam K. Garg, Shiv Patel, Wonjun Yim, Jesse Jokerst, Daniel L. Chao

## Abstract

**Importance:** The global COVID-19 pandemic, which began in late 2019, has resulted in a renewed focus on the importance of personal protective equipment (PPE) and other interventions to decrease the spread of infectious diseases. While several ophthalmology organizations have recently released guidance on appropriate PPE for surgical procedures and ophthalmology clinics, there is limited experimental evidence demonstrating the efficacy of the various interventions that have been suggested. In this study, we evaluate high-risk aspects of the slit-lamp exam and the effect of various PPE interventions.

**Objective:** To determine the effectiveness of surgical masks, slit lamp shields, and gloves in decreasing the spread of large droplets in an ophthalmology clinical setting and evaluating high-risk sources of contamination.

**Design:** Experimental cough simulation using a fluorescent surrogate of respiratory droplets during an ophthalmic slit lamp examination.

**Setting:** Single-center

**Participants:** Patient Simulation

**Main outcomes and measure(s):** Presence of fluorescent particles in the air near or on slit lamp and simulated slit lamp examiner.

**Results:** Simulated coughing without a mask or slit lamp shield resulted in widespread dispersion of fluorescent droplets on the model slit lamp examination. Coughing with a mask resulted in the most significant decrease in fluorescent droplets, however, particles were still seen to escape from the top of the mask. Coughing with the slit lamp shield alone blocked the majority of forward particle dispersion; however significant distributions of respiratory droplets were found on the slit lamp joystick as well as table. Coughing with both mask and slit lamp shield resulted in the least dispersion on to the simulated examiner. Scanning electron microscopy demonstrated particle sizes of 3-100μm.

**Conclusion and relevance:** Masking has the greatest effect in limiting spread of respiratory droplets, while slit lamp shields, and gloves also contribute to limiting exposure to respiratory droplets from SARS-CoV-2 during slit lamp examination.

**Key Points:** *Question:* How effective are certain personal protective equipment and interventions in the clinic in preventing respiratory droplet spread?

*Findings:* Simulating patient coughing during the slit lamp exam demonstrated that masking appeared to be the most effective intervention to prevent spread of respiratory droplets. Slit lamp shields and gloves were also effective in decreasing exposure to respiratory droplets.

*Meaning:* This study confirms the importance of masking, slit lamp shields, as well as gloves as important interventions to limit exposure to respiratory droplets from respiratory viruses such as SARS-CoV-2.

## Introduction

The COVID-19 global pandemic has had a profound effect on clinical care around the world. As a result of the outbreak, many ophthalmology clinics temporarily halted elective clinical visits and surgical procedures. Furthermore, the pandemic has resulted in fervent discussion regarding best practices to limit the spread of infectious diseases in the clinical setting. This disease, like many other upper respiratory infections, is highly transmissible via respiratory droplets, with recent reports suggesting airborne transmission of the can also occur^1–3^.

Several independent ophthalmology organizations, including the American Academy of Ophthalmology (AAO), have released guidelines regarding the resumption of clinical care as well as recommendations on appropriate personal protective equipment (PPE) that should be used when performing providing patient care. AAO guidelines recommend surgical masks for patients, masks and eye protection for providers, and slit lamp breath shields^4^. However, there has been limited evidence regarding the efficacy of these interventions. As clinical activities resume, there is a need for robust data to inform use and efficacy of PPE in the clinic. Recent work compared several commercially available slit lamp shields for degree of respiratory droplet spread protection, providing necessary evidence for best clinical practice^1^. However, several open questions remain for the development of best-practice, evidence-based guidelines for PPE during the ophthalmic exam. In this study, we develop a patient cough simulator to evaluate high-risk areas of respiratory droplet contamination during a slit lamp examination (Figure 1).

**Figure 1:**
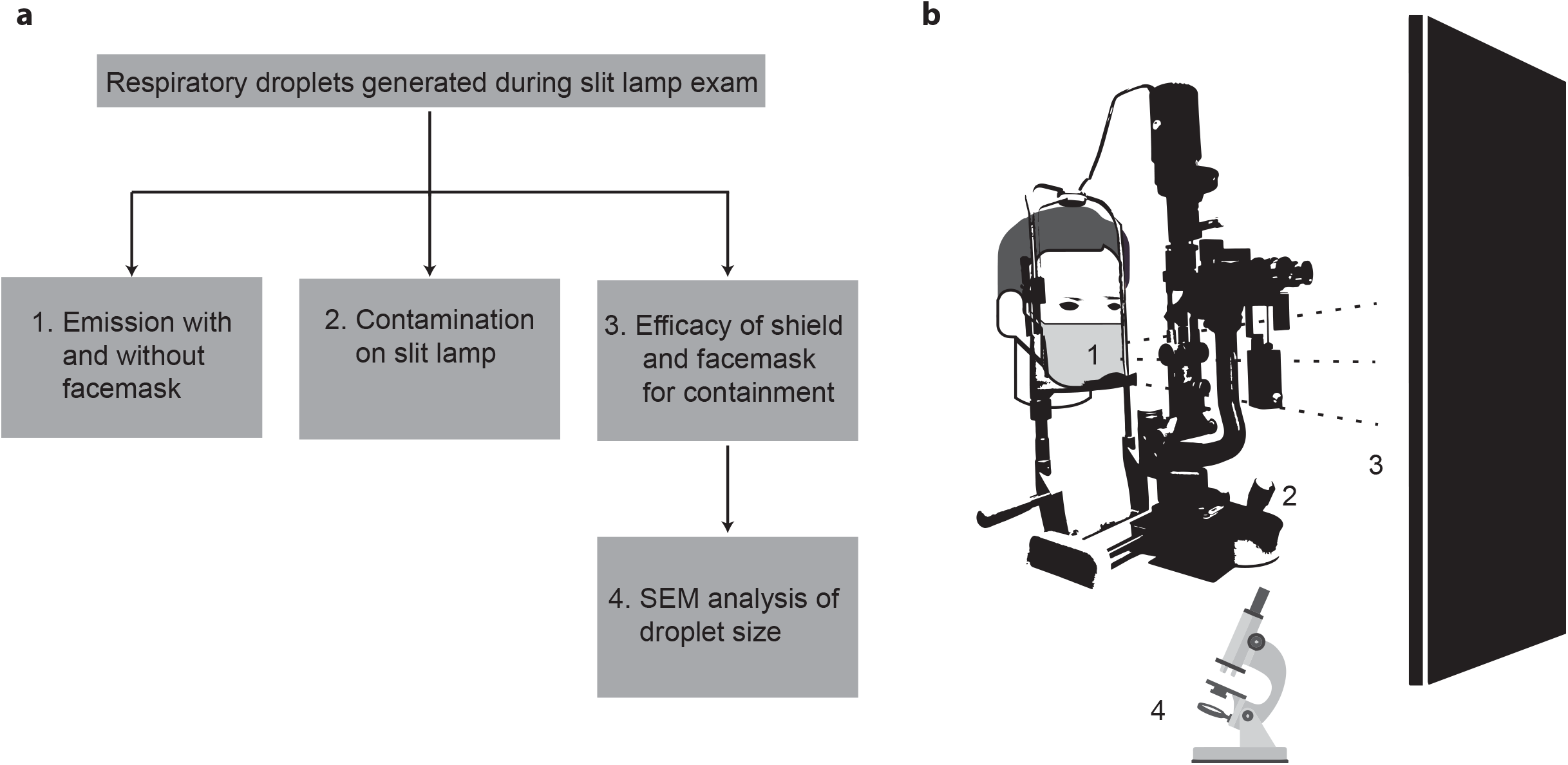
Overview of experimental strategy. **a**. 4 aspects of respiratory risk during the slit lamp exam were investigated: 1) effect of surgical masks on droplet emission 2) droplet contamination on the slit lamp and table 3) droplets reaching the examiner 4) microscopic analysis of droplets. **b**. Depiction of slit lamp experimental design.

## Methods

### Coughing simulator

GloGerm MIST, a fluorescent surrogate of respiratory droplets used in previous simulation studies, was used as a simulator for patient coughing^5^. GloGerm MIST was sprayed using the included pressurized spray canister, and the resulting mist was visualized using a handheld ultraviolet flashlight (Escolite 51 LED 395 nM Ultraviolet Blacklight Detector). The pressurized canister was placed inside of a 3D printed head to simulate a patient exam. Details of 3D printing are available upon request.

For some trials, a blue surgical mask (3M Standard Procedure Mask) was placed over the mouth and nose. All simulated coughs were 1 second in duration. GloGerm speed was estimated at 0.7m/s (Supplemental Movie 2). To capture respiratory droplets reaching the slit lamp examiner, a 78cm x 114cm wooden black board was placed at the level of the oculars and imaged between trials (Figure 1b).

### Slow motion droplet video recording and respiratory droplet visualization

All photographs and videos were taken using a tripod-mounted Sony A7III digital camera. The blacklight was placed in a stationary position and video was recorded at 120fps at 1080p resolution. Photographs were taken in a dark room with a 5-second exposure. All post-capture editing was completed in Adobe Photoshop (photos) or Adobe Premiere Pro 2020 (videos) with all changes in contrast and brightness identical in all trials. Two still frames were exported for representative images in Figure 2. For visualization of differences between still images, photos were auto-aligned, the difference was calculated between images, then the resulting difference was inverted and thresholded.

**Figure 2:**
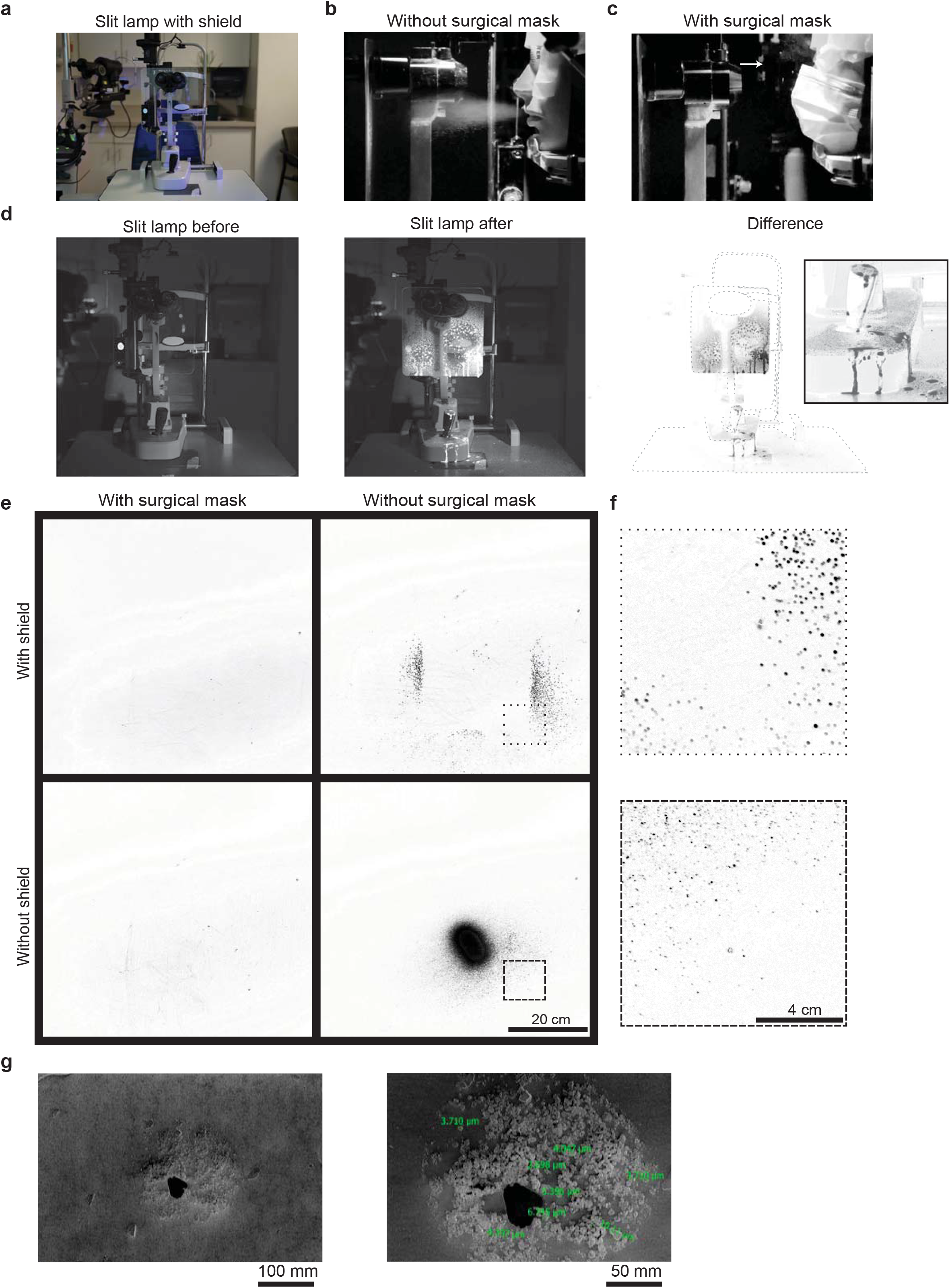
Efficacy of PPE on respiratory droplet spread. **a**. Photograph of slit lamp with slit lamp shield used for all experiments. **b**. Still image from Supplemental Movie 1 demonstrating forward moving particles during experimental trial with unmasked simulated patient. **c**. Same as b, with masked simulated patient. Arrows demonstrate respiratory particles ejected upward. **d**. Before (left), after (middle), and subtracted (right) image of slit lamp from experimental trial with unmasked simulated patient. Inlay of slit lamp joystick (right) demonstrating significant contamination of joystick. **e**. Subtracted images of each experimental condition. **f**. Zoomed-in regions from e, depicted in e with dashed outlines. **g**. Scanning electron microscopy (SEM) images of simulated respiratory particles (left). Individual measurements, in green, of fluorescent GloGerm particles within larger respiratory droplet (right).

### SEM Quantification

Carbon tape (Nisshin EM) was used to capture GloGerm droplets. A scanning electron microscope (SEM; FEI Apreo) was used to examine the size and morphology of droplets at an acceleration voltage of 1kV.

## Results

Slow motion video recordings were taken of GloGerm respiratory droplets ejected from the 3D printed head to simulate a forceful cough. Identical recordings with a surgical mask over the patient’s mouth demonstrate the drastic reduction in forward moving droplets (Figure 2b-c, Supplemental Movie 1). Of note, respiratory particles were seen ejected upwards past the nares with the facemask present (Figure 2c, white arrow).

The slit lamp was next imaged before and after a simulated cough without a mask but with a slit lamp shield. When the difference between the images were calculated, we observed a majority of the respiratory droplets contained by the shield. However, we identified significant contamination on the joystick as well as the slit lamp table (Figure 2d).

We next assessed how well the surgical mask and slit lamp shield prevented respiratory droplets from reaching the examiner. We sprayed GloGerm through the simulated patient with or without a mask, and with or without a slit lamp shield present. When we compared images of the board before and after the simulated cough, we were able to identify the pattern of droplets reaching the examiner (Figure 2e). When no protection was in place, a dense core of droplets can be seen. A shield alone was able to block most of the central droplets, however peripheral particles can still be seen (Figure 2f). Minimal droplets were identified when the face mask was present. Together, these data suggest a combination of a surgical mask and a slit lamp shield can block a vast majority of forward-moving respiratory droplets.

Finally, we determined the microscopic architecture of the individual droplets (Figure 2f) seen in the previous experiment. Using SEM, we identified 3-10μm droplet nuclei contained within one ∼150μm respiratory droplet (Figure 2g).

## Conclusion

In this study, we characterized the risk to ophthalmologists from patients coughing through a simulated patient slit lamp examination. Using simulated respiratory droplets, we found the most effective intervention for containing spread was masking for both patient and physician. When combined with a slit lamp shield, a majority of respiratory droplets are blocked from reaching the examiner. We also identify the slit lamp joystick and examination table as a high-risk area for contamination given its location under the shield, suggesting that gloves may prevent physician contact with droplets. Our simulator is likely an underestimation of particle spread, as GloGerm primarily forms droplets of 100μm, which is much larger than most respiratory (>5-10μm) or airborne particles (<5μm)^6^.

Respiratory particles were observed escaping through the top of the mask lateral to the nose, consistent with previous findings^7^, posing a risk when providers are close to the patient’s face such as during intravitreal injections or when using a direct ophthalmoscope. While further studies are needed to fully evaluate the risk of virus transmission during close encounters, our data suggest that use of eye protection is prudent.

Recent work suggests that surgical masks may be sufficient for reducing emission of viral particles greater than 5μm, but less effective below that range^8^. Our SEM evaluation of respiratory droplets reaching the provider identified droplets ranging from 3-100μm, suggesting patient facemask alone is not sufficient to prevent all risk of respiratory or airborne contamination.

There are many limitations to this study. First GloGerm MIST may not be an exact representation of respiratory secretions during a cough. While we are able to capture larger particles, many of the droplet nuclei under 1μm are not detected by our fluorescent methodology. In addition, we only tested one size slit lamp shield and one type of mask as a proof-of-concept. Further work characterizing the efficacy of different commercially available masks and shields is necessary for a complete understanding of the risks associated with clinic visits.

As ophthalmologic clinic appointments start to resume, additional data are needed to provide best evidence-based guidelines for appropriate PPE in the clinic. Our cough simulation experiments lend support for universal masking, slit lamp shields, as well as gloves to limit exposure to potential SARS-CoV-2 during the ophthalmic exam.

## Data Availability

All data is available upon reasonable request.

## Acknowledgements

D.L.C. was funded by NIH/NEI K08EY030510 and an unrestricted department grant for Research to Prevent Blindness. J.J was funded by University of California Tobacco Related-Disease Research Program (TRDRP) under Emergency COVID-19 Research Seed Funding award number R00RG2515, the NIH under grants DP2 HL137187 and R21 AG065776, and the NSF under grant #1845683.

## Author Contributions

This study was conceived by S.H.S, A.K.G., S.P., and D.L.C. Experimental design was done by S.H.S., A.K.G., S.P., W.Y., J.J., and D.L.C. Acquisition of data was by S.H.S., A.K.G., S.P., and W.Y. Analysis and interpretation of data were done by S.H.S., A.K.G., S.P., W.Y., J.J., and D.L.C. Drafting of the paper was done by S.H.S. and A.K.G. Revision of the paper was done by S.H.S., A.K.G., S.P., and D.L.C.

---

**Supplemental Movie 1: High-speed capture of simulated cough with and without surgical mask**

Three trials of GloGerm cough simulation with and without a surgical mask were recorded at 120fps and slowed to 0.1x speed. With a mask, a cloud of respiratory droplets is visible escaping upwards lateral to the nose (arrow).

**Supplemental Movie 2: Simulated respiratory droplet velocity**

A simulated cough was recorded at 120fps next to a measuring tape. White lines were placed at the frontline of respiratory droplets in 4 consecutive frames to estimate average velocity. The video was then slowed down for visualization.

## Supplemental Methods

### 3D printing parameters

An open-source 3D head file was modified using Blender first by cutting along two planes in order to reconfigure the dimensions. Then, a boolean difference was performed using an 8.0cm x 8.0cm x 19.7cm cylinder to make a cylindrical hole parallel to the head. A hole orthogonal to the previous cylinder to provide an opening at the lips for spraying GloGerm. The head was printed on an Ultimaker s5 with PLA, 0.2mm layer height, and 25% infill.

